# Dual-platform plasma proteogenomics prioritizes GSTA1 and ARG1 as druggable cross-domain targets in cardiovascular-kidney-metabolic syndrome

**DOI:** 10.64898/2026.11.14.26368123

**Authors:** Youmian Lan, Chujia Lin, Hu Liu, Houshi Zhou, Bingying Xie, Jiahao Chen, Heng Gao, Qiongbing Zheng, Yongsong Chen

## Abstract

**Background:** Cardiovascular-kidney-metabolic (CKM) syndrome reflects shared pathophysiology across metabolic, renal and cardiovascular systems, yet circulating proteins with causal effects spanning multiple CKM domains remain unclear. We aimed to identify and prioritize cross-domain candidate proteins using an integrative, dual-platform plasma proteogenomic framework.

**Methods:** We combined genetic causal inference, statistical colocalization, single-cell expression mapping, and structure-based druggability assessment. *Cis*-protein quantitative trait loci (*cis*-pQTL) instruments were derived from two independent plasma proteomic platforms, UKB-PPP (2,030 proteins with valid cis instruments) and deCODE (1,640 proteins). Genetic causal inference (two-sample Mendelian randomization) was performed against seven CKM-related outcomes across the metabolic (type 2 diabetes [T2D], body mass index [BMI]), renal (estimated glomerular filtration rate [eGFR], chronic kidney disease) and cardiovascular (coronary artery disease, heart failure, stroke) domains. Proteins showing false discovery rate (FDR)-significant associations in at least two CKM domains independently on both platforms were prioritized, and cross-platform directional concordance was evaluated as a robustness measure. Prioritized proteins were followed by colocalization, and characterization using single-cell expression, disease-association and druggability resources. Structural and ligand-efficiency analyses were performed for the leading candidate using its co-crystallized inhibitor complex.

**Results:** Proteome-wide analysis yielded 9,734 (UKB-PPP) and 8,837 (deCODE) protein-outcome estimates, with 77.9% two-platform directional concordance. Cross-domain prioritization identified six proteins (ARG1, GSTA1, INHBB, ITIH1, NCAN, SHBG), all acting across the metabolic-renal axis. Colocalization provided strong support for GSTA1 with eGFR (posterior probability of a shared causal variant 0.97) and ARG1 with BMI (0.93). Genetically higher GSTA1 was associated with lower eGFR and higher T2D risk, with kidney expression confined to proximal tubule cells. Genetically higher ARG1 was associated with lower BMI and lower T2D risk, consistent with hepatocyte-restricted expression. Both proteins were small-molecule tractable. For GSTA1, the inhibitor-bound structure (PDB 1GSF) showed a deep, enclosed H-site, and three chemotypes with previously reported low-micromolar potency against recombinant hGSTA1-1 (ethacrynic acid and the dietary flavonols fisetin and myricetin) all exceeded the lead-like ligand-efficiency threshold (0.30 kcal mol^-1^ per heavy atom).

**Conclusions:** Using dual-platform proteogenomics, we identify genetically supported cross-metabolic-renal candidate proteins in CKM syndrome, with GSTA1 and ARG1 emerging as the most robust, pharmacologically tractable candidates. These findings nominate actionable targets and mechanistic entry points for CKM syndrome.

## Introduction

Cardiovascular-kidney-metabolic (CKM) syndrome, formally defined by the American Heart Association in 2023, describes the interconnected pathophysiology linking metabolic risk factors, chronic kidney disease (CKD) and cardiovascular disease[1–3]. Rather than progressing as isolated conditions, metabolic dysfunction, renal impairment and cardiovascular injury frequently co-occur and reinforce one another, conferring substantially elevated morbidity and mortality[4, 5]. More than one in three US adults have multiple CKM risk factors, and CKM stages are highly prevalent in the general population[6, 7]. This integrated framework has shifted emphasis from single-organ risk factors toward shared upstream mechanisms operating across organ systems, and toward unified risk prediction and management[8, 9], creating a pressing need to identify molecular drivers common to multiple CKM domains that could serve as unifying therapeutic targets.

The circulating proteome offers a uniquely informative window onto such shared mechanisms, since plasma proteins integrate signals from multiple tissues and are themselves accessible druggable targets, with genetic support substantially increasing the probability of successful drug development[10, 11]. Recent large-scale plasma proteogenomic studies, including the UK Biobank Pharma Proteomics Project (UKB-PPP)[12] and deCODE[13], have mapped protein quantitative trait loci (pQTLs) for thousands of proteins[14]. When a *cis*-protein quantitative trait loci (*cis*-pQTL) is used as a genetic instrument, Mendelian randomization (MR) can infer the causal effect of a protein’s genetically predicted level on disease, mitigating confounding and reverse causation that limit observational proteomic associations[15]. Coupled with colocalization[16], this *cis*-MR strategy has become a powerful approach for nominating causal, tractable protein targets[17, 18].

To date, however, proteome-wide MR studies have largely focused on individual diseases or isolated organ systems, such as diabetic kidney disease[19] or coronary artery disease (CAD)[20, 21]. Because CKM syndrome is driven by mechanisms shared across organ systems, proteins with causal effects spanning multiple CKM domains are especially attractive therapeutic candidates: a single such target could, in principle, be modulated to benefit the metabolic, renal and cardiovascular arms simultaneously. Identifying these proteins, however, requires the simultaneous, multi-outcome interrogation that single-disease proteome-wide MR is not designed to provide. Findings derived from a single proteomic platform can be influenced by aptamer- or antibody-specific artefacts and residual confounding[22], and single-variant cis colocalization is inherently conservative[23], underscoring the value of cross-platform replication. For example, single-outcome proteome-wide MR studies have implicated GSTA1 and INHBC in type 2 diabetes (T2D)[24] and multiple proteins have been implicated in estimated glomerular filtration rate (eGFR) regulation[25], but each within a single CKM domain.

Here we integrate two independent plasma proteomic platforms (UKB-PPP and deCODE) with GWAS of seven outcomes spanning the metabolic, renal and cardiovascular domains to systematically identify cross-domain causal proteins in CKM syndrome. Using a proteome-wide MR framework that combines platform-specific cross-domain screening, cross-platform concordance assessment and colocalization, we prioritize a focused set of genetically supported proteins acting across the metabolic-renal axis and identify GSTA1 and ARG1 as robust, druggable candidates with concordant tissue expression and mechanistic plausibility. Our findings nominate actionable molecular targets and mechanistic entry points for CKM syndrome.

## Methods

### Data Availability

All data analyzed in this study are publicly available summary-level datasets, detailed in Table 1. The crystal structure is available from the Protein Data Bank (accession 1GSF). Analysis scripts and derived result tables are available from the corresponding author upon reasonable request.

**Table 1.** Detailed information on data sources used in the Mendelian randomization analyses.

| Traits | Consortium | Cases | Controls | Sample size | Year | Population | PubMed (PMID) | Website / source |
|---|---|---|---|---|---|---|---|---|
| T2D<br>(metabolic) | DIAMANTE/DIAGRAM | 80,154 | 853,816 | 933,970 | 2022 | European | 35551307 | diagram-consortium.org/downloads.html |
| BMI<br>(metabolic) | GIANT | – | – | 322,154 | 2015 | European | 25673413 | portals.broadinstitute.org/collaboration/giant/ |
| eGFR<br>(renal) | CKDGen | – | – | 567,460 | 2019 | European | 31152163 | ckdgen.imbi.uni-freiburg.de/datasets/Wuttke_2019 |
| CKD<br>(renal) | CKDGen | 41,395 | 439,303 | 480,698 | 2019 | European | 31152163 | ckdgen.imbi.uni-freiburg.de/datasets/Wuttke_2019 |
| CAD<br>(cardiovascular) | CARDIoGRAMplusC4D | 60,801 | 123,504 | 184,305 | 2015 | European | 26343387 | cardiogramplusc4d.org/data-downloads/ |
| HF<br>(cardiovascular) | HERMES | 40,805 | 542,362 | 583,167 | 2020 | European | 31919418 | kp4cd.org/node/283 |
| Any stroke<br>(cardiovascular) | GIGASTROKE | 73,652 | 1,234,808 | 1,308,460 | 2022 | European | 36180795 | ebi.ac.uk/gwas/studies/GCST90104539 |
| Plasma proteome<br>(exposure) | UKB-PPP | – | – | 34,557* | 2023 | European | 37794186 | Primary publication, Supplementary Table 10 |
| Plasma proteome<br>(exposure) | deCODE | – | – | 35,559 | 2021 | Icelandic | 34857953 | Primary publication, Supplementary Table 2 |

**Abbreviations:** T2D, type 2 diabetes; BMI, body mass index; eGFR, estimated glomerular filtration rate; CKD, chronic kidney disease; CAD, coronary artery disease; HF, heart failure; \* UKB-PPP profiled 2,923 proteins in 54,219 participants overal; the pQTL discovery summary statistics used for instrument selection were generated in 34,557 European-ancestry participants.

### Study design

We performed a dual-platform, two-sample proteome-wide MR study to identify circulating proteins with genetically supported effects across multiple CKM domains, followed by colocalization and multi-omic characterization. The workflow comprised instrument construction, discovery MR, cross-domain prioritization, sensitivity analyses, colocalization, and expression and druggability annotation **(Fig. 1)**, and adhered to the STROBE-MR guidelines[26]. Analyses used summary-level GWAS data from populations of European ancestry. The contributing studies had received approval from their corresponding ethics committees and participants had provided informed consent. The Ethics Committee of the First Affiliated Hospital of Shantou University Medical College determined that additional ethics approval was not required for this secondary analysis of publicly available summary-level data.

**Figure 1.**
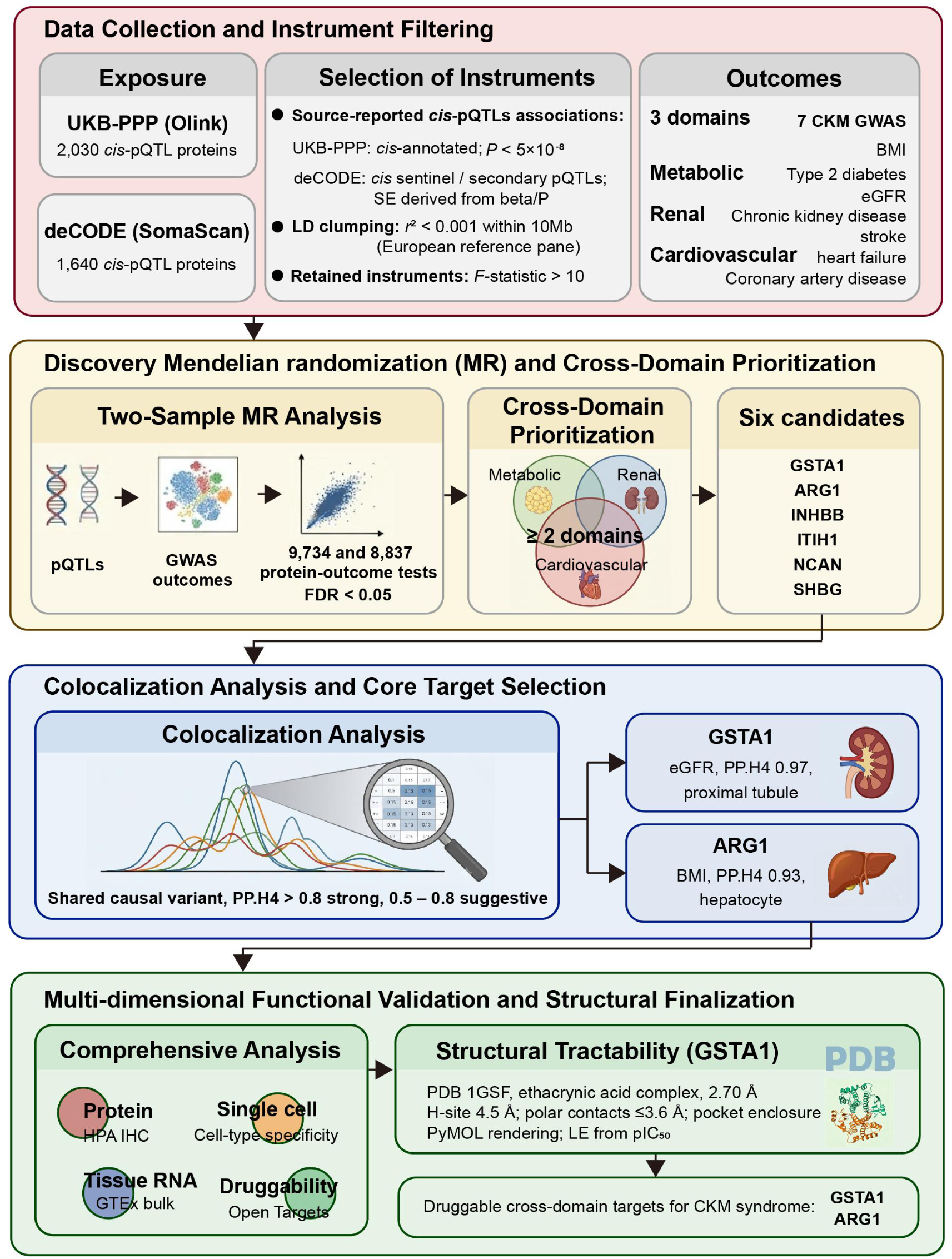
Study design. Dual-platform *cis*-pQTL instruments derived from UKB-PPP (Olink) and deCODE (SomaScan) were tested for causal associations with seven CKM-relevant outcomes spanning metabolic, renal, and cardiovascular domains using two-sample Mendelian randomization, followed by cross-domain prioritization, colocalization analysis, and multi-omic functional characterization.

MR inference relies on three core instrumental-variable assumptions: relevance, independence from confounders, and exclusion restriction. Relevance was supported by restricting analyses to strong *cis*-pQTL instruments (*F* > 10) after linkage-disequilibrium (LD) clumping. Ancestry-matched GWAS datasets and minimization of exposure-outcome sample overlap were used to reduce potential violations of independence, whereas restriction to *cis*-pQTLs, cross-platform replication and colocalization were used to reduce concerns regarding horizontal pleiotropy and linkage-disequilibrium confounding. The study was not preregistered.

### Exposure data and instrument construction

*Cis*-pQTL instruments were derived from two independent plasma proteomic resources: UKB-PPP (Olink Explore)[12] and deCODE (SomaScan)[13]. UKB-PPP profiled 2,923 unique proteins in 54,219 participants overall, with the pQTL discovery summary statistics used for instrument selection were generated in 34,557 European-ancestry participants, whereas deCODE measured 4,907 aptamers in 35,559 Icelandic participants. For UKB-PPP, source-reported cis variants from the primary association table were used, with genome-wide significance defined as *P* < 5×10^-8^. For deCODE, source-reported *cis* sentinel and secondary pQTL associations reported in the original proteomic GWAS were used, and standard errors were reconstructed from the reported unadjusted beta and *P* value. Instruments with an *F*-statistic [*F* = (beta/SE)²] ≤ 10 were excluded to avoid weak-instrument bias. Independent instruments were obtained by LD clumping (*r*² < 0.001, 10,000 kb) using a European reference panel. After quality control and clumping, 2,030 UKB-PPP proteins and 1,640 deCODE proteins with valid cis instruments were included in MR. No a priori power calculation was performed; instrument strength was evaluated using *F*-statistics.

### Outcome data

Seven GWAS of CKM-related outcomes were selected across three domains: metabolic — T2D (DIAMANTE, European)[27] and body mass index (BMI) (GIANT)[28], renal — eGFR and CKD (CKDGen)[29], and cardiovascular — CAD (CARDIoGRAMplusC4D)[30], heart failure (HF) (HERMES)[31] and any stroke (GIGASTROKE)[32]. Trait definitions followed those of the cited source consortia. Full sample sizes, exact downloaded datasets, accession information and repositories are provided in **Table 1**. Where available, outcome GWAS excluding the UKB were used (e.g., HERMES minus-UKB for HF). Any residual sample overlap therefore concerns only the UKB-PPP-based analyses, whereas deCODE was drawn from an independent Icelandic cohort. Datasets were harmonized to genome build GRCh37. For T2D, BMI, eGFR, CKD, CAD and HF, variants were matched primarily by rsID, multi-allelic and indel variants with ambiguous mapping were excluded. The GIGASTROKE any-stroke release was indexed by GRCh37 chromosome: position rather than rsID. Accordingly, UKB-PPP instruments were matched to stroke using their GRCh37 coordinates, whereas deCODE standard rsIDs were mapped to GRCh37 coordinates using dbSNP155 (SNPlocs.Hsapiens.dbSNP155.GRCh37) before chromosome: position matching. Effect and non-effect alleles were aligned using TwoSampleMR, and palindromic variants with ambiguous allele frequencies were excluded. Protein-outcome pairs with no valid instrument after harmonization were not analyzed.

### Discovery Mendelian randomization

For each protein-outcome pair on each platform, two-sample MR was performed using the TwoSampleMR framework[33]. The Wald ratio was used for single-instrument proteins and inverse-variance weighted (IVW) MR for proteins with multiple instruments[15]. Effect estimates were expressed as odds ratios (binary outcomes) or beta coefficients (continuous outcomes) per one standard-deviation increase in genetically predicted protein level. Multiple testing was controlled by the Benjamini-Hochberg false discovery rate (FDR) within each outcome, with FDR < 0.05 considered significant. Published GWAS summary estimates were used without additional covariate adjustment, covariate sets therefore followed the respective source studies rather than being harmonized across samples.

### Cross-domain prioritization

Each outcome was mapped to one of three CKM domains (metabolic, renal, cardiovascular). Within each proteomic platform separately, a protein was classified as cross-domain if it showed FDR-significant associations in at least two distinct CKM domains. The intersection of the platform-specific cross-domain sets was retained. Cross-platform directional concordance of shared protein-outcome associations was then quantified as a robustness metric; the candidate-defining significant associations were directionally concordant between platforms. Retained proteins were carried forward for colocalization and multi-omic characterization.

### Sensitivity analyses

Directionality was assessed using the MR Steiger test for candidate-defining protein-outcome associations[34]. Cochran’s Q was additionally assessed for multi-variant candidate associations where estimable; MR-Egger pleiotropy testing was not estimable because no prioritized association had three or more instruments. Cross-platform concordance and colocalization were used as complementary robustness checks. Colocalization was used to distinguish shared causal signals from confounding by linkage disequilibrium between distinct nearby signals and was not interpreted as proof of the exclusion-restriction assumption.

### Colocalization

To distinguish causal effects from confounding by linkage disequilibrium, we performed Bayesian colocalization (coloc.abf) between *cis*-pQTL and outcome GWAS signals within the ±500 kb *cis* region of each candidate gene[16]. Full summary statistics were used, with SNPs matched by rsID across datasets. Effect-allele frequencies for the proteomic data were obtained from the corresponding annotation file. Default priors were applied. Colocalization was assessed using the posterior probabilities of a shared causal variant (PP.H4) and of two distinct causal variants (PP.H3). A PP.H4 > 0.8 was considered strong evidence of colocalization and 0.5 – 0.8 suggestive, whereas a high PP.H3 with low PP.H4 indicated distinct signals.

### Expression and druggability annotation

Leading candidates were characterized using publicly available resources. Tissue and single-cell expression were examined in the Human Protein Atlas (https://www.proteinatlas.org/) and GTEx (https://www.gtexportal.org/). Disease associations and target tractability were obtained from Open Targets (https://platform.opentargets.org/). These analyses contextualized the mechanistic plausibility and therapeutic potential of the prioritized proteins.

### Structural and pharmacological characterization of GSTA1

GSTA1 tractability was assessed from the crystal structure of human GSTA1-1 bound to ethacrynic acid (PDB 1GSF, 2.70 Å)[35], using the chain A/B biological homodimer (REMARK 350) with waters removed. Open-source PyMOL (v2.5.10) was used for structural calculations and rendering. H-site residues were defined as chain A residues within 4.5 Å of the co-crystallized ligand, and polar contacts as ligand–protein N/O pairs ≤ 3.6 Å. Pocket enclosure was quantified as the fraction of ligand solvent-accessible surface buried on binding (probe radius 1.4 Å). Ligand efficiency was calculated as LE = 1.37 × pAffinity / N_heavy[36] from published in vitro potencies against recombinant hGSTA1-1.

### Statistical analysis

Effect estimates are expressed as odds ratios (binary outcomes) or beta coefficients (continuous outcomes) per 1-SD increase in genetically predicted protein level, and are reported with standard errors and 95% confidence intervals; unless otherwise stated, the measure of variance is the standard error. Multiple testing was controlled using the Benjamini-Hochberg false discovery rate (FDR) within each outcome, with FDR < 0.05 considered significant. All statistical tests were two-sided. All analyses were conducted in R software (v4.5.0) using TwoSampleMR (v0.6.14), ieugwasr (v1.1.0), coloc (v5.2.3), and the Bioconductor SNPlocs.Hsapiens.dbSNP155.GRCh37 resource for rsID-to-GRCh37 coordinate mapping in the stroke analysis. Structural analyses were performed using PyMOL (v2.5.10). Figures were generated using ggplot2 (v4.0.1) and related packages.

## Results

### Study design and instrument construction

To systematically identify circulating proteins with causal effects spanning multiple domains of CKM syndrome, we implemented a dual-platform proteome-wide MR framework **(Fig. 1)**. *Cis*-pQTL instruments were derived independently from two large plasma proteomic resources based on orthogonal assay technologies: UKB-PPP (Olink; 2,030 proteins with valid *cis* instruments) and deCODE (SomaScan; 1,640 proteins). These were tested against seven GWAS of CKM-related outcomes spanning the metabolic (T2D, BMI), renal (eGFR, CKD) and cardiovascular (CAD, HF, stroke) domains, selected to minimize sample overlap with the exposure cohorts **(Table 1)**. All retained instruments were strong, with minimum *F*-statistics of 45.37 in UKB-PPP and 20.83 in deCODE (all *F* > 10).

### Proteome-wide MR across CKM outcomes

Two-sample MR of each protein against each outcome yielded 9,734 and 8,837 protein-outcome estimates for UKB-PPP and deCODE, respectively, across the seven CKM outcomes. After Benjamini-Hochberg correction within each outcome (FDR < 0.05), the number of significant proteins was highest for renal traits (eGFR: 128 [UKB] and 120 [deCODE]) and metabolic traits (T2D: 77 and 66). For the corrected any-stroke analysis, 13 UKB-PPP and 6 deCODE associations reached FDR < 0.05; HF yielded 3 and 0, respectively **(Fig. 2A)**. Volcano plots show the distribution of associations and the positions of the eventual cross-domain candidates for all seven outcomes **(Fig. 2B, C)**.

**Figure 2.**
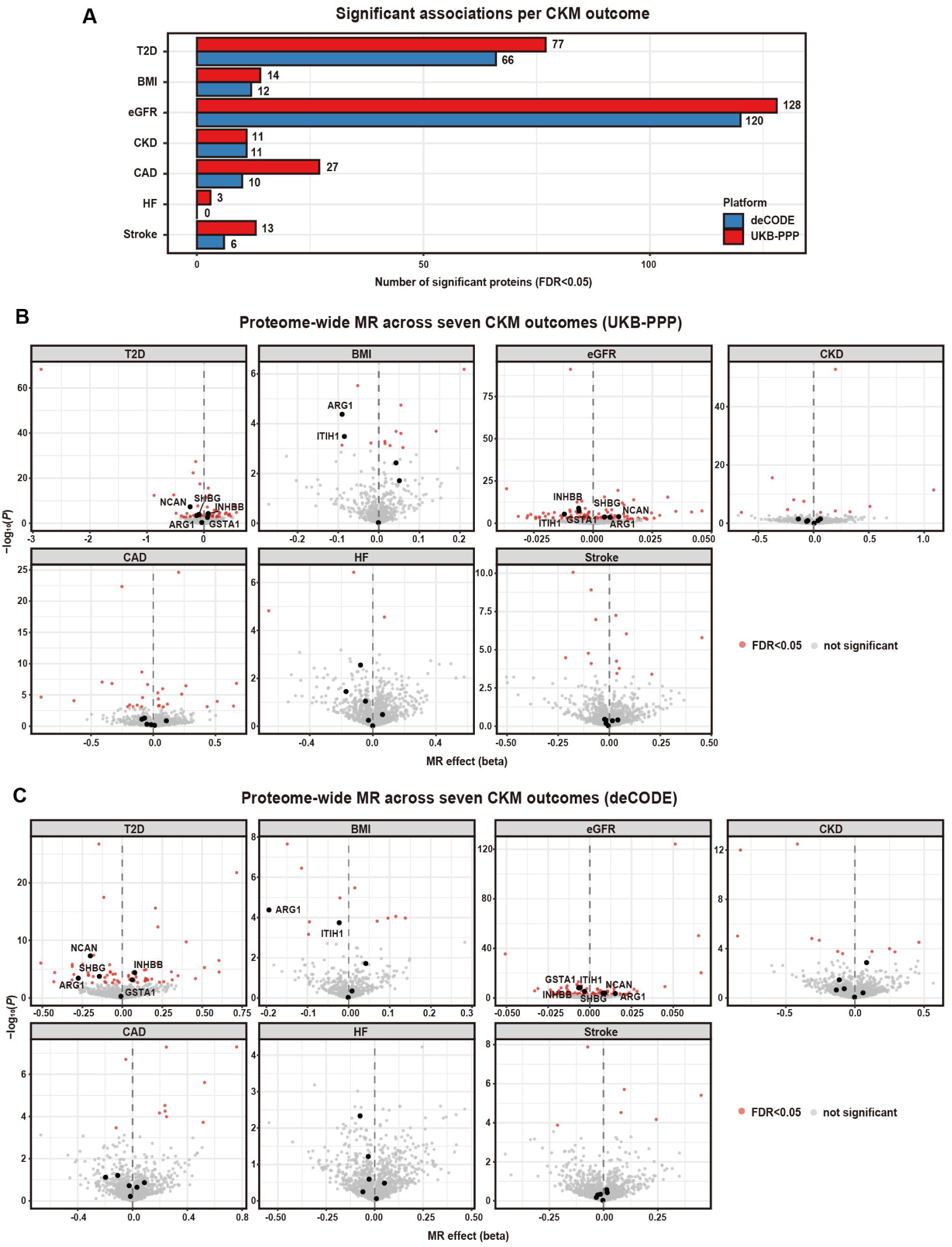
Proteome-wide MR across CKM outcomes. **A.** Number of FDR-significant protein–outcome associations (FDR < 0.05) for each CKM outcome, by platform (blue, deCODE; red, UKB-PPP). **B.** Volcano plots of MR effect estimates (beta) versus −log₁₀(*P*) for each outcome using UKB-PPP instruments. Red, FDR < 0.05; gray, ns. Cross-domain candidates labeled where significant. **C.** As in (B), using deCODE instruments.

### Concordance across two independent platforms

UKB-PPP and deCODE differ in source populations and assay chemistries, making cross-platform concordance an important robustness check. Among 3,831 protein-outcome pairs tested on both platforms, MR estimates were directionally concordant in 77.9% of cases, with a Pearson correlation of *r* = 0.55 for the effect estimates **(Fig. 3A)**. Fifty-five proteins reached FDR significance on both platforms across the seven outcomes **(Fig. 3B)**.

**Figure 3.**
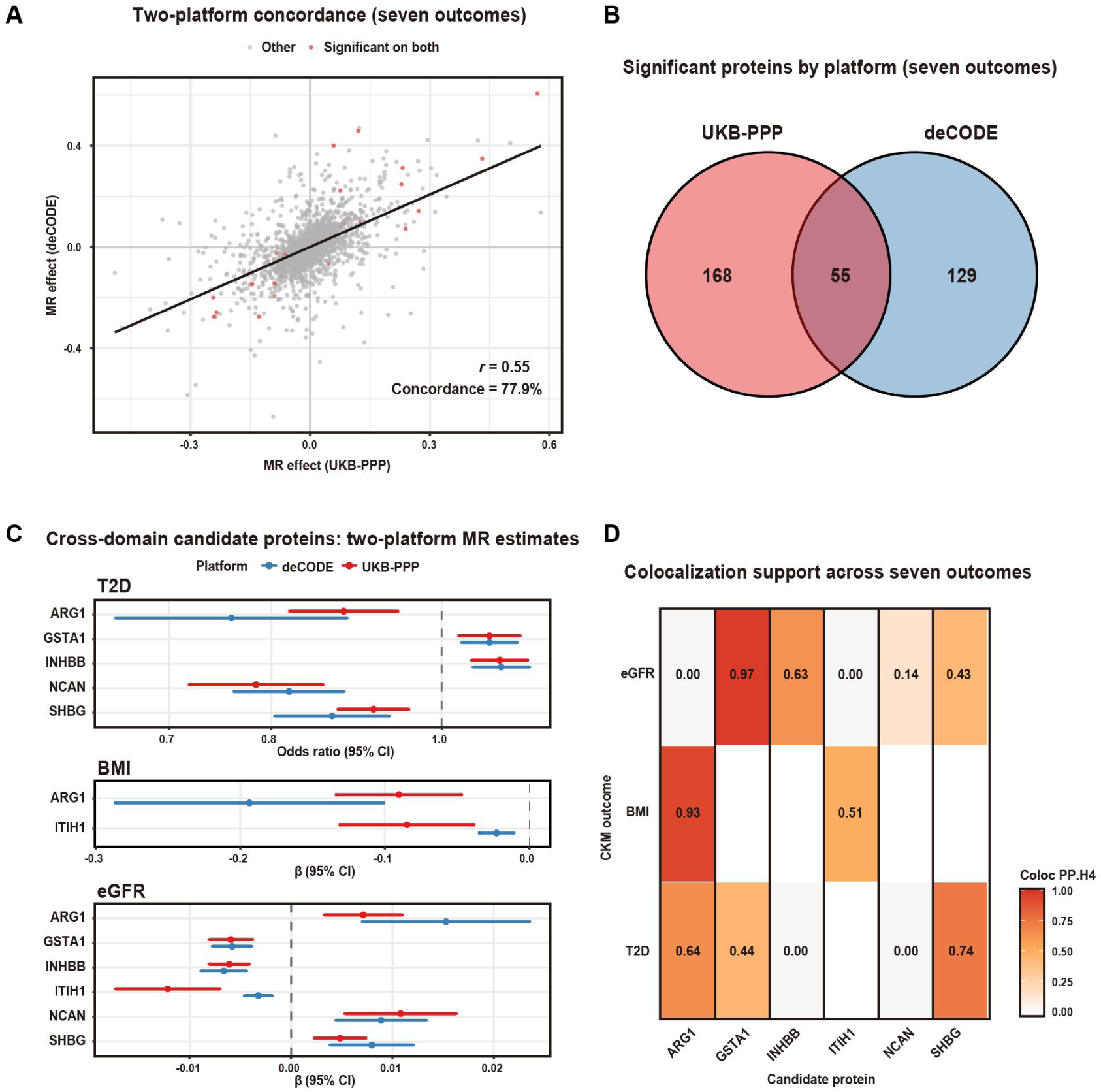
Two-platform robustness and cross-domain candidates. **A.** Concordance of MR effects estimates between UKB-PPP and deCODE across all tested proteins (seven outcomes combined). Red, significant on both platforms (FDR < 0.05); gray, other. **B.** Overlap of FDR-significant proteins identified by each platform. **C.** Two-platform MR effect estimates for the six cross-domain candidates across candidate-defining outcomes, binary T2D effects are shown as log ORs and continuous BMI/eGFR effects as beta coefficients, with 95% confidence intervals. **D.** Colocalization support (PP.H4) between candidate proteins and CKM outcomes.

### Cross-domain prioritization identifies six candidate proteins

Given the shared pathophysiology underlying CKM syndrome, we prioritized proteins showing cross-domain associations on both platforms. Applying the platform-specific criterion of FDR-significant associations in at least two CKM domains and then intersecting the two platform-specific sets yielded six cross-domain candidates: ARG1, GSTA1, INHBB, ITIH1, NCAN and SHBG. All six acted across the metabolic-renal axis, and none met the cross-domain criterion through a cardiovascular outcome.

Candidate-defining significant associations had concordant directions between platforms. For example, ARG1 was associated with lower T2D risk (OR 0.879 [UKB-PPP] and 0.759 [deCODE]), whereas GSTA1 was associated with lower eGFR (beta −0.0059 and −0.0058, respectively), as summarized in **Table 2** and **Fig. 3C**. The harmonized variant-level *cis*-pQTL and corresponding outcome associations underlying these candidate-defining MR estimates are provided in **Table S1**.

**Table 2.** Two-platform Mendelian randomization estimates and colocalization evidence for the six cross-domain candidate proteins.

| Protein | Outcome | UKB-PPP estimate (95% CI) | DeCODE estimate (95% CI) | Dir. | Coloc PP.H4 | Evidence |
|---|---|---|---|---|---|---|
| GSTA1 | eGFR | -0.0059 (-0.0081, -0.0038) | -0.0058 (-0.0077, -0.0039) | 1.00 | 0.97 | Strong |
| GSTA1 | T2D | 1.0640 (1.022, 1.108) | 1.0640 (1.027, 1.103) | 1.00 | 0.44 | MR only |
| ARG1 | BMI | -0.0903 (-0.1335, -0.0471) | -0.1936 (-0.2863, -0.1010) | 1.00 | 0.93 | Strong |
| ARG1 | T2D | 0.8790 (0.819, 0.944) | 0.7590 (0.653, 0.883) | 1.00 | 0.64 | Suggestive |
| ARG1 | eGFR | 0.0071 (0.0033, 0.0110) | 0.0153 (0.0071, 0.0235) | 1.00 | 0.00 | Not coloc |
| SHBG | T2D | 0.9140 (0.873, 0.957) | 0.8660 (0.804, 0.934) | 0.83 | 0.74 | Suggestive |
| SHBG | eGFR | 0.0048 (0.0023, 0.0074) | 0.0080 (0.0039, 0.0121) | 0.83 | 0.43 | MR only |
| INHBB | eGFR | -0.0061 (-0.0081, -0.0041) | -0.0066 (-0.0089, -0.0044) | 1.00 | 0.63 | Suggestive |
| INHBB | T2D | 1.0780 (1.040, 1.118) | 1.0810 (1.041, 1.121) | 1.00 | 0.00 | Not coloc |
| ITIH1 | BMI | -0.0847 (-0.1309, -0.0385) | -0.0227 (-0.0347, -0.0108) | 0.86 | 0.51 | Suggestive |
| ITIH1 | eGFR | -0.0121 (-0.0173, -0.0070) | -0.0032 (-0.0046, -0.0018) | 0.86 | 0.00 | Not coloc |
| NCAN | T2D | 0.7840 (0.719, 0.856) | 0.8190 (0.762, 0.880) | 1.00 | 0.00 | Not coloc |
| NCAN | eGFR | 0.0108 (0.0053, 0.0163) | 0.0089 (0.0044, 0.0134) | 1.00 | 0.14 | MR only |

For T2D, effect estimates are odds ratios (ORs) with 95% confidence intervals per one standard-deviation increase in genetically predicted plasma protein level. For the continuous outcomes BMI and eGFR, estimates are beta coefficients with 95% confidence intervals on the source GWAS scale. Dir. is the fraction of all shared protein-outcome pairs for a given protein with concordant effect direction across platforms. PP.H4 is the posterior probability that the pQTL and outcome association share a causal variant. **Abbreviations:** T2D, type 2 diabetes; BMI, body mass index; eGFR, estimated glomerular filtration rate.

MR Steiger analyses supported the hypothesized protein-to-outcome direction for all 13 candidate-defining associations on both proteomic platforms (26/26 platform-specific tests). Most prioritized associations were instrumented by a single *cis*-pQTL. Among the two deCODE INHBB associations with two instruments, Cochran’s Q showed no evidence of between-variant heterogeneity for T2D (*P* = 0.783) or eGFR (*P* = 0.262); MR-Egger pleiotropy testing was not estimable because no prioritized association contained three or more instruments.

### Colocalization refines causal candidates

Colocalization between *cis*-pQTL and outcome GWAS signals was assessed for each candidate. GSTA1–eGFR (PP.H4 = 0.97) and ARG1–BMI (PP.H4 = 0.93) showed strong colocalization, marking these as the most robustly supported pairs. Suggestive colocalization (0.5 < PP.H4 < 0.8) was observed for SHBG–T2D (0.74), ARG1–T2D (0.64), INHBB–eGFR (0.63) and ITIH1–BMI (0.51), whereas NCAN showed weak colocalization overall (T2D: PP.H4 = 0.00; eGFR: PP.H4 = 0.14). Several pairs with significant MR estimates lacked colocalization support and instead showed high PP.H3 (e.g., INHBB-T2D), consistent with distinct neighboring signals **(Fig. 3D**, **Fig. 4)**.

**Figure 4.**
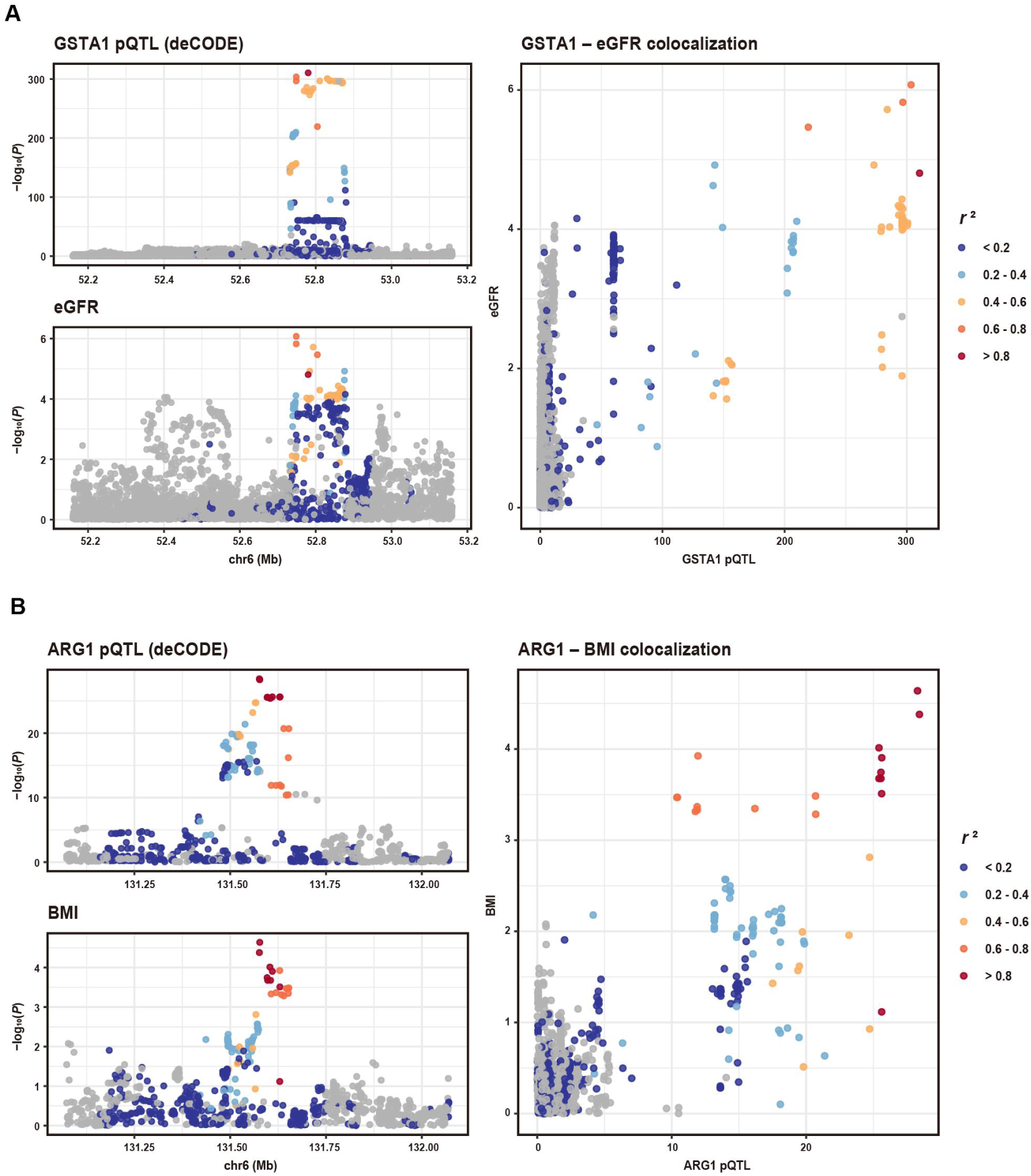
Colocalization (LocusCompare) for leading candidates: GSTA1–eGFR and ARG1–BMI. **A.** Regional association plots for GSTA1 pQTL (deCODE, top) and eGFR (bottom) at the GSTA1 locus, and corresponding LocusCompare scatter plot (right) showing colocalization between the two signals. Points colored by LD (*r*²) with the lead variant. **B.** As in (A), for ARG1 pQTL (deCODE) and BMI at the ARG1 locus.

### Tissue expression and druggability of the leading proteins

Given their strong colocalization support, we further characterized GSTA1 and ARG1 using multi-omic resources to assess tissue specificity and druggability. GSTA1 protein showed moderate staining in renal tubular cells but was absent from glomeruli, and single-cell RNA data localized its expression mainly to proximal tubule cells **(Fig. 5A, 5B)**. ARG1 was strongly expressed in hepatocytes at both protein and single-cell RNA levels **(Fig. 5A, 5C)**. At the tissue level, GTEx data showed broad GSTA1 expression across adrenal gland, liver, and kidney cortex **(Fig. 5D)**, whereas ARG1 expression was almost exclusively liver-restricted, matching its single-cell profile **(Fig. 5E)**. Disease-association data (Open Targets) linked GSTA1 to metabolic, hepatic-steatotic and renal phenotypes, stroke and T2D, and ARG1 to T2D, obesity, metabolic syndrome and stroke, corroborating their involvement across the CKM spectrum **(Fig. 5F, 5G)**. Both proteins were assessed as small-molecule tractable: GSTA1 belongs to a druggable protein family with a high-quality binding pocket and ligand-bound structures, while ARG1 has characterized small-molecule ligands and resolved structures **(Fig. 5H, 5I)**.

**Figure 5.**
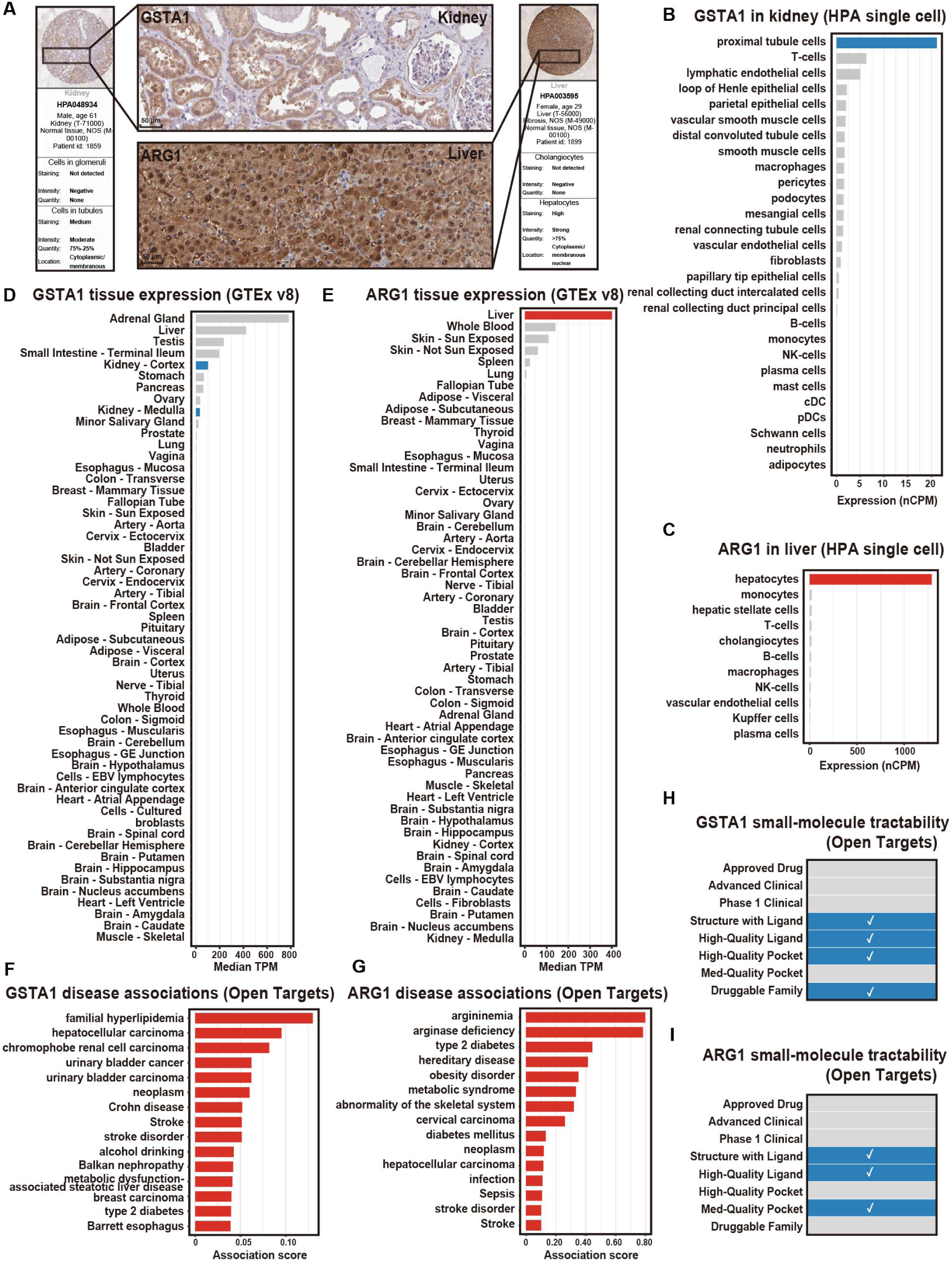
Tissue expression and druggability of GSTA1 and ARG1. **A.** Representative immunohistochemistry of GSTA1 in kidney (top; antibody HPA048934) and ARG1 in liver (bottom; antibody HPA003595) from the Human Protein Atlas. Scale bars, 50 µm. **B.** GSTA1 single-cell RNA expression in kidney (HPA), shown as normalized counts per million (nCPM); enriched in proximal tubule cells (blue). **C.** ARG1 single-cell RNA expression in liver (HPA), shown as normalized counts per million (nCPM); enriched in hepatocytes (red). **D.** GSTA1 expression across GTEx v8 tissues, shown as median transcripts per million (TPM); kidney cortex highlighted in blue. **E.** ARG1 expression across GTEx v8 tissues, shown as median transcripts per million (TPM); liver highlighted in red. **F.** Top GSTA1 disease associations (Open Targets), ranked by association score. **G.** Top ARG1 disease associations (Open Targets), ranked by association score. **H.** GSTA1 small-molecule tractability (Open Targets); check marks indicate satisfied criteria. **I.** ARG1 small-molecule tractability (Open Targets); check marks indicate satisfied criteria.

### Structural basis and pharmacological tractability of GSTA1

Open Targets tractability scoring **(Fig. 5H)** flags GSTA1 as small-molecule tractable, without indicating a binding site or mechanism, so we examined its resolved structure and the pharmacology of known inhibitors. GSTA1-1 is an obligate homodimer, and its electrophile-binding H-site is defined experimentally by the inhibitor ethacrynic acid, a clinically approved diuretic and a classical GST inhibitor (PDB 1GSF; **Fig. 6A**). The ligand is 89.3% buried on binding (solvent-accessible surface 456.1 → 48.9 Å²), indicating a deep, enclosed pocket rather than a shallow surface site. The 14 contacting residues are 79% hydrophobic (11/14), with polar anchoring from polar bonds between the ligand carboxylate and the catalytic Tyr9 hydroxyl (2.64 Å) and the Val55 backbone carbonyl (2.66 Å) **(Fig. 6B, Table S2)**.

**Figure 6.**
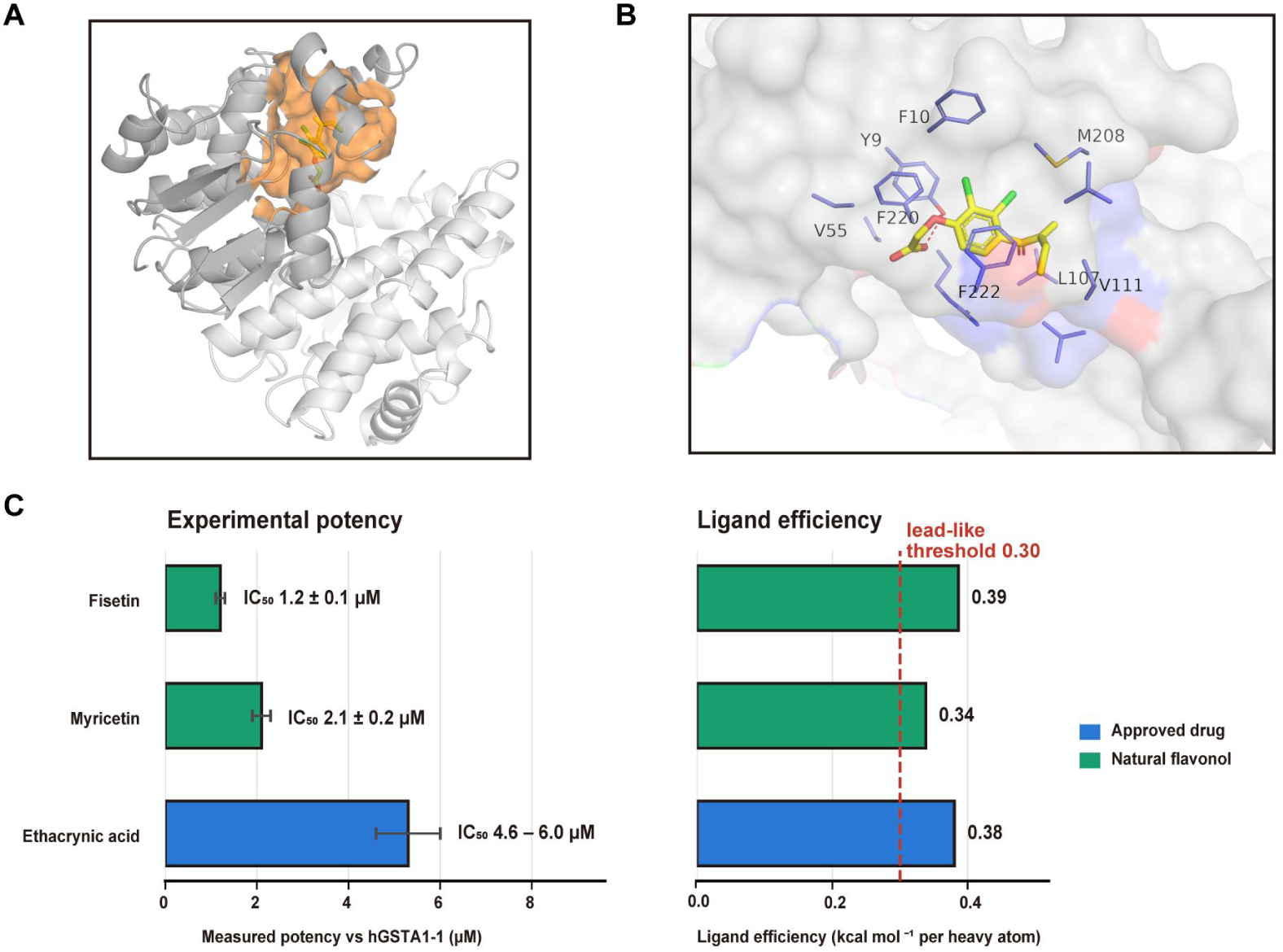
Structural basis and ligand efficiency of GSTA1 inhibition. **A.** Crystal structure of the GSTA1-1 homodimer with ethacrynic acid bound in the H-site (PDB 1GSF). Residues within the binding pocket are highlighted in orange. **B.** Close-up of the H-site pocket. Ethacrynic acid (yellow) is enclosed by contacting residues (labeled), with the molecular surface colored by hydrophobicity. **C.** Left: measured potency (IC_50_, µM) against recombinant hGSTA1-1 for ethacrynic acid, fisetin, and myricetin. Right: ligand efficiency (kcal mol⁻¹ per heavy atom); the dashed line marks the lead-like threshold of 0.30. Bars colored by compound class (blue, approved drug; green, natural flavonol). Where reported, error bars indicate SD; ethacrynic acid is shown as the published potency range.

Three structurally distinct chemotypes have been reported to inhibit purified recombinant hGSTA1-1 at low-micromolar potency: ethacrynic acid (half-maximal inhibitory concentration [IC_50_] 4.6 – 6.0μM)[37], and the dietary flavonols fisetin (IC_50_ 1.2 ± 0.1μM)[38] and myricetin (IC_50_ 2.1 ± 0.2 μM)[39]. All three exceed the conventional lead-like ligand-efficiency threshold of 0.30 kcal mol^-1^ per heavy atom (≈ 0.380, 0.386 and 0.338 respectively; **Fig. 6C**), indicating that the potency reflects efficient binding rather than molecular size. GSTA1 therefore satisfies standard structural and ligand-efficiency criteria for tractability, with existing inhibitors spanning an approved drug and low-toxicity natural-product scaffolds.

## Discussion

In this dual-platform proteome-wide MR study, we systematically searched for circulating proteins with genetically supported effects spanning multiple domains of CKM syndrome. Six cross-domain candidates emerged from the intersection of the two platform-specific screens, all acting across the metabolic-renal axis. GSTA1 and ARG1 showed the strongest convergent support: colocalization was strong for GSTA1 with eGFR and ARG1 with BMI, and both are small-molecule tractable.

Across both platforms, genetically higher GSTA1 was associated with lower eGFR and higher risk of T2D, defining a consistent renal-metabolic risk profile. The eGFR signal was the most robustly supported component, being significant on both platforms and anchored by strong colocalization; the proximal-tubule-restricted renal expression of GSTA1 further points toward a tubular origin. GSTA1 is an alpha-class glutathione S-transferase abundant in proximal tubular epithelium and released into the urine upon tubular stress, and is an established biomarker of proximal-tubule injury[40]. A higher genetically predicted circulating level may therefore mark greater tubular vulnerability, consistent with the inverse association with eGFR. Because *cis*-pQTL MR indexes genetically proxied circulating abundance rather than tissue enzymatic activity, these results do not establish that direct pharmacological inhibition of GSTA1 would reproduce the MR effect. Notably, the GSTA1–T2D association has been independently reported in prior proteome-wide MR[24, 41], and our two-platform analysis extends the signal to the renal domain within a unified cross-domain framework. Beyond the genetic evidence, the structural features of GSTA1 support its tractability: it assembles into an obligate homodimer, and the H-site of each subunit is well defined, having been resolved in complex with the inhibitor ethacrynic acid[35], a validated GSTA1 inhibitor[37]. Importantly, this pocket is not restricted to a single chemotype — the structurally unrelated dietary flavonols fisetin[38] and myricetin[39] also inhibit recombinant hGSTA1-1 with IC₅₀ values in the low-micromolar range.

Genetically higher ARG1 was associated with lower BMI and lower T2D risk on both platforms, providing a coherent protective signal in the metabolic arm of CKM syndrome; the BMI association was strongly colocalized, and the T2D association showed suggestive colocalization. This direction is independently concordant with a prior dual-platform proteome-wide MR of T2D[42]. ARG1 encodes arginase 1, expressed in hepatocytes and in alternatively activated (M2) macrophages, where it is a canonical anti-inflammatory marker. The protective metabolic association is biologically consistent with ARG1’s association with M2 macrophage polarization and reduced adipose inflammation in obesity[43, 44]. Conversely, mitochondrial arginase 2 promotes macrophage inflammation, insulin resistance and atherogenesis, suggesting that the two isoforms may act differently[45, 46]. ARG1 was also associated with higher eGFR on both platforms, but that pair did not colocalize (PP.H4 = 0.00); we therefore do not regard the renal association as comparably robust. As noted for GSTA1, *cis*-pQTL MR proxies circulating abundance rather than enzymatic activity, and the protective direction therefore argues for isoform- and tissue-specific experimental validation before therapeutic conclusions are drawn.

Together, GSTA1 — a risk-associated protein of the renal-metabolic axis, and ARG1 — a protective protein of the metabolic arm — illustrate how a cross-domain framework can resolve distinct and even opposing target hypotheses. Both were supported by two orthogonal proteomic platforms and by at least one strongly colocalized protein-outcome pair. Their therapeutically favorable directions are target-specific: the genetic results are consistent with lowering the GSTA1-associated tubular-injury axis, whereas the ARG1 findings, together with the opposing role of ARG2, argue against assuming that broad arginase inhibition would be beneficial. Such directional resolution is difficult to obtain from single-outcome or single-platform designs, and it is relevant to the emerging paradigm of cross-domain pharmacotherapy in CKM syndrome, exemplified by agents such as SGLT2 inhibitors and GLP-1 receptor agonists that act across the metabolic and renal domains[47]. This directional information is a strength of MR-based target prioritization, but MR reflects lifelong genetically proxied differences in protein abundance rather than the effects of short-term therapeutic intervention[48]. Functional studies are therefore required to determine whether modulating GSTA1 activity or the ARG1-associated axis can reproduce these genetically predicted effects in a tissue- and isoform-specific manner. This cross-domain, cross-platform design distinguishes our study from prior proteome-wide MR work that typically focuses on a single outcome[19–21], and replication across two independent proteomic technologies reduces the risk of platform-specific false positives[22]. Nevertheless, a few limitations should be noted. Our analyses were limited to populations of European ancestry, which restricts generalizability. Although FDR-significant associations were observed in individual cardiovascular outcomes, including the corrected stroke analysis, no prioritized protein satisfied the cross-domain criterion through a cardiovascular outcome. This should not be interpreted as evidence that shared cardiovascular mechanisms are absent; trait-specific genetic architecture, instrument availability and the statistical power of currently available GWAS, particularly for heart failure, may all contribute. Circulating protein levels may not reflect activity within the target tissue, and single-variant colocalization can be conservative when multiple causal variants co-occur in a region[23]. Requiring agreement across two independent proteomic technologies[12, 13] together with colocalization support limits, but does not eliminate, platform-specific artefacts, horizontal pleiotropy and linkage-related confounding. Finally, MR estimates capture the effect of lifelong genetically predicted protein levels, which may not correspond to what would be observed with short-term pharmacological intervention.

## Conclusion

Using a dual-platform, cross-domain proteome-wide MR framework, we identify GSTA1 and ARG1 as genetically supported candidate proteins linking the metabolic and renal arms of CKM syndrome. Their strongest signals are supported by convergent MR, colocalization and tissue-expression evidence, and the two proteins carry opposite therapeutic directions. Experimental studies are warranted to determine whether these genetically informed target hypotheses can be translated into therapeutic strategies.

## Acknowledgments

The authors thank Prof. Stanley Li Lin from Shantou University Medical College for English language editing.

## Author contributions

**Youmian Lan**: Conceptualization, Methodology, Investigation, Formal analysis, Writing - original draft, Writing - review & editing, Funding acquisition, Project administration; **Chujia Lin**: Conceptualization,Writing - review & editing, Funding acquisition; **Hu Liu**: Methodology, Investigation, Formal analysis, Writing - original draft; **Houshi Zhou**: Conceptualization, Methodology, Investigation, Formal analysis; **Bingying Xie**: Conceptualization, Investigation, Formal analysis; **Jiahao Chen**: Methodology, Investigation, Formal analysis; **Heng Gao**: Methodology, Investigation, Formal analysis; **Qiongbing Zheng**: Methodology, Investigation, Formal analysis, Writing - original draft, Writing - review & editing, Project administration; **Yongsong Chen**: Conceptualization, Writing - review & editing, Funding acquisition, Resources, Supervision. All authors read and approved the final manuscript.

## Abbreviations

BMI: body mass index
CAD: coronary artery disease
*cis*-pQTL: *cis*-protein quantitative trait loci
CKD: chronic kidney disease
CKM: cardiovascular-kidney-metabolic
eGFR: estimated glomerular filtration rate
HF: heart failure
FDR: false discovery rate
IC_50_: half-maximal inhibitory concentration
MR: Mendelian randomization
PP.H3: posterior probability of hypothesis 3
PP.H4: posterior probability of hypothesis 4
pQTLs: protein quantitative trait loci
T2D: type 2 diabetes
UKB-PPP: UK Biobank Pharma Proteomics Project

